# The dynamics of arterial pressure itself predict intraoperative hypotension beyond its current value: an interpretable additive model validated in 3,069 external patients under a selection-bias-resistant protocol

**DOI:** 10.64898/2026.08.26.26361468

**Authors:** Ricardo Oyarzun, Pablo Hernández

## Abstract

**Background:** Whether predictors of intraoperative hypotension (IOH) add information beyond the mean arterial pressure (MAP) already displayed on the monitor is contested: selection bias in common evaluation designs inflates apparent performance, and the field has called for comparisons against simple MAP-based references under bias-resistant protocols. Existing predictors also depend on proprietary waveform analysis or pulse-contour monitors, restricting both deployment and external validation.

**Methods:** Using 807 non-cardiac surgery patients from the open VitalDB database, we derived an *additive* gradient boosting model (one split per tree: a learned shape function per variable, no interactions) from three variables computable from an arterial line alone: current MAP, its drift from the patient’s own 20-minute baseline, and the growth of its rolling variance (critical slowing down). Evaluation used patient-level 5-fold cross-validation under a strict protocol - exclusion of the 65-75 mmHg grey zone and of all samples already hypotensive at prediction time - with MAP alone (same learner class) as comparator. The frozen model was then validated, without any refitting, on an independent cohort from another continent (MOVER, University of California Irvine) following a pre-registered plan sealed before external data access.

**Results:** In development the pressure-only model reached AUROC 0.907 vs. 0.884 for MAP alone (ΔAUROC +0.023, 95% CI +0.017 to +0.029) at 5 min, with +0.031 and +0.032 at 10 and 15 min, and good calibration (Brier skill +0.418 vs. prevalence). In external validation on 3 069 patients (442 194 samples, 1-minute charting, event prevalence 5.8%), the advantage not only transferred but was larger than in development: AUROC 0.696 vs. 0.638, ΔAUROC +0.058 (95% CI +0.051 to +0.064), meeting both pre-registered gates. Discrimination transferred; calibration did not (external Brier skill -0.014), requiring local recalibration. In the unrestricted scenario, where samples already at threshold are retained, the advantage collapsed (+0.007), reproducing the selection effect this paper documents. A secondary model adding pulse-contour cardiac output and stroke volume variation improved development discrimination further (ΔAUROC +0.035) but could be externally validated in only 39 patients, because those signals are rarely recorded.

**Conclusions:** The dynamics of arterial pressure itself — drift from a patient-specific baseline and variance growth — carry predictive information beyond its current value, in a fully interpretable additive model that requires only an arterial line, no waveform access and no proprietary hardware. The advantage is confirmed in a pre-registered frozen-model external validation of over three thousand patients, and is largest at coarse recording cadence, where instantaneous pressure is least informative.

## 1 Introduction

Intraoperative hypotension is associated with acute kidney injury, myocardial injury and death, with risk rising as mean arterial pressure falls below 65 mmHg [1], and its reported incidence varies from 5% to 99% depending on the definition applied [2]. Machine-learning predictors, most prominently the waveform-based Hypotension Prediction Index [3], report high discrimination; however, re-analyses have shown that common evaluation designs inflate apparent performance through selection bias, and that much of the apparent skill is attributable to the current MAP value itself [4], [7]. The field’s response has been methodological: predictors should demonstrate added value over simple MAP-based references [5] — including linear extrapolation of MAP (LepMAP) — under protocols that exclude the threshold grey zone [6].

This paper takes that challenge literally. We ask two questions. First, do low-frequency haemodynamic trends — variables available from any pulse-contour monitor at 20-second cadence, without waveform access — add predictive information beyond current MAP when evaluated under the strictest available protocol? Second, if they do, can that information be extracted by a model that remains fully interpretable?

Our starting point is dynamical-systems theory: circulatory decompensation as a critical transition preceded by generic early-warning signals — variance growth, phase-space expansion, drift from equilibrium [10] — a paradigm already proposed for acute transitions in clinical medicine [12]. In earlier work we combined three such precursors in a fixed-weight linear index. Here we show that the linear functional form, not the choice of variables, was the bottleneck: an additive model — one learned shape function per variable, no interaction terms — recovers essentially all extractable information while remaining as inspectable as a regression table, and its learned shapes exhibit precisely the threshold behaviour that bifurcation theory predicts.

## 2 Methods

### 2.1 Data source and cohort

We used VitalDB, an open database of 6 388 surgical cases from Seoul National University Hospital [9]. We included all non-cardiac surgery cases with pulse-contour cardiac output monitoring (Vigileo/FloTrac: 262 patients; EV1000/HemoSphere: 545 patients; total 807) and at least one hour of concurrent MAP, CO and SVV recording, yielding 616 703 twenty-second observations (3426 patient-hours). Invasive MAP came from the arterial line (Solar8000/ART _MBP, with EV1000/AR _MBP as fallback); heart rate from ECG; BIS where available (default 50 when absent). IOH was defined as MAP <65 mmHg sustained for ≥60 s (three consecutive samples). The first 15 min of each case were excluded (induction); the following 20 min defined the patient-specific baseline.

### 2.2 Variables

Nine strictly causal variables entered the model, all computable in real time from backward-looking windows: the five monitored channels (MAP, CO, SVV, HR, BIS) and four dynamical signals computed over a 10-min rolling window: (i) *MAP drift*, the difference between windowed MAP and the patient s baseline mean, in units of the baseline SD; (ii) *phase-space volume*, the log-determinant of the rolling covariance of [MAP, CO, SVV]; (iii) *critical slowing down*, the ratio of windowed MAP variance to baseline MAP variance; (iv) *fixed-point distance*, the distance in normalised state space to the equilibrium of per-patient sparse polynomial dynamics identified on the baseline segment. Savitzky-Golay derivatives were computed with centred windows during preprocessing and were therefore *excluded* from all predictive models: we verified empirically that a centred 9-sample window reacts up to 80 s before a step change, i.e. it filters future information into the present.

### 2.3 Evaluation protocol

Reporting follows TRIPOD+AI [13]. Following the recommendations of Jacquet-Lagréze et al. [6] and the selection-bias analysis of Enevoldsen and Vistisen [4], the primary analysis used a **strict scenario**: all post-baseline samples, excluding (a) samples with current MAP in the 65-75 mmHg grey one and (b) samples already meeting the IOH definition at prediction time. Sensitivity analyses report the no-exclusion and 65-70 mmHg variants. Labels *y*(*t*) indicate sustained IOH onset within the horizon (5, 10, 15 min). All models were evaluated with patient-level stratified 5-fold cross-validation (no patient in both training and validation); z-scoring parameters were fitted on training folds only. Confidence intervals are patient-level bootstrap (2 000 replicates). Comparison thresholds (ΔAUROC ≥ 0.02 with CI excluding zero) were frozen before any model comparison was run.

### 2.4 Models

The primary model is additive gradient boosting (`HistGradientBoostingClassifier, max_leaf_nodes=` 2, learning rate 0.06, up to 800 trees with early stopping): each tree makes a single split, so the ensemble is an additive combination of one-dimensional step functions — a shape function per variable, with no interactions, directly visualisable. Comparators: (i) MAP alone under an unrestricted boosting learner of the same family (a stronger baseline than logistic MAP, since it captures the threshold non-linearity); (ii) LepMAP, linear extrapolation of MAP from a 10-min window to the horizon [6]; (iii) linear logistic regression on the nine variables; (iv) spline GAM; (v) unrestricted boosting with interactions. Probabilities were calibrated by isotonic regression fitted within training folds. Clinical utility was assessed by decision-curve analysis 11J. he final model was refitted on the full cohort and frozen (with its exact variable specification) before any external data access.

### 2.5 Formal verification of the points score (Lean 4)

The printed score sheet (Supplement S1) is a quantisation of the additive model, and we verified the fidelity of that quantisation formally, in the Lean proof assistant, using exact integer arithmetic (fixed point, 10^−6^ log-odds) and no external mathematical libraries: every proof is checked by the Lean kernel. Three theorems are machine-checked over the frozen derivation-cohort model: (T1) for each variable, the difference between the exact shape function and 0.05*×* its integer points is bounded by a computed per-variable constant, for *every* possible input value; ( 2) globally, the model s log-odds and 0.05*×* the printed score differ by at most *B* = 0.617 log-odds for every possible patient state (triangle inequality over 1); ( 3) as a corollary, any two states whose model log-odds differ by more than 2*B* = 1.235 can never be mis-ranked by the printed score. *B* is dominated by the sparse low-MAP tail; over the bulk of the population the discrepancy is far smaller. These theorems certify the *functional* fidelity of the deployed artefact to the validated model — they assert nothing about clinical accuracy, which rests on the cross-validated and confirmatory results above. The Lean development is part of the public repository.

## 3 Results

### 3.1 Primary model: arterial pressure dynamics alone

**Table 1.** Pressure-only additive model (MAP, drift from the patient s own baseline, critical slowing down) versus MAP alone, in development and in frozen-model external validation. Strict scenario. Patient-level bootstrap CIs (2 000 replicates).

| Cohort | Horizon | $n$ patients | Pressure model | MAP alone | ΔAUROC (95% CI) |
| --- | --- | --- | --- | --- | --- |
| VitalDB (development) | 5 min | 807 | 0.907 | 0.884 | +0.023 (+0.017 to +0.029) |
| VitalDB (development) | 10 min | 807 | 0.839 | 0.808 | +0.031 (+0.023 to +0.039) |
| VitalDB (development) | 15 min | 807 | 0.802 | 0.769 | +0.032 (+0.023 to +0.041) |
| MOVER (external) | 5 min | 3069 | 0.696 | 0.638 | +0.058 (+0.051 to +0.064) |
| MOVER (external) | 10 min | 3069 | 0.668 | 0.609 | +0.059 — |
| MOVER (external) | 15 min | 3069 | 0.656 | 0.600 | +0.056 — |

Three variables computable from an arterial line reached AUROC 0.90 at 5 min in development versus 0.88 for MAP alone (ΔAUROC +0.023, CI +0.01 to +0.029), rising to +0.032 at 15 min — that is, at longer hori ons the pressure-only model retains almost the entire advantage of the full nine-variable model (Section 3.2). Calibration was good (Brier skill +0.418).

In external validation the frozen model was applied unchanged to 3 069 MOVER patients (442 194 samples). The advantage over MAP was *larger* than in development: +0.058 (CI +0.051 to +0.064 ), and stable across horizons (+0.059 and +0.056). Both pre-registered gates were met. We read this magnification as a cadence effect rather than a superiority claim: MOVER charts at one-minute resolution, where the instantaneous MAP value is a weaker predictor (AUROC 0.638 vs. 0.88 in development), whereas drift and variance are window averages and degrade far less. The practical implication is that the model helps most exactly where monitoring is coarsest.

Two honest qualifications. Calibration did *not* transfer (external Brier skill -0.014 ): the model s probabilities require local recalibration before any threshold-based use, as expected given a different event prevalence and cadence. And in the unrestricted scenario — retaining samples already at or below threshold — the advantage collapses to +0.007, reproducing externally the selection effect documented below.

### 3.2 Model ladder: where the information lies

**Table 2.** AUROC under the strict scenario. The step from the linear to the additive row isolates non-linearity as the missing ingredient of fixed-weight indices; the step from additive to unrestricted (negligible or negative) shows interactions add nothing generalisable.

| Model | AUROC 5 min | AUROC 10 min | AUROC 15 min |
| --- | --- | --- | --- |
| MAP alone (gradient boosting) | 0.884 | 0.808 | 0.769 |
| Published 4-term index | 0.749 | 0.717 | 0.696 |
| Linear logistic, 9 variables | 0.894 | 0.839 | 0.808 |
| GAM (cubic splines) | 0.914 | 0.856 | 0.824 |
| **Additive model (this work)** | 0.919 | 0.859 | 0.825 |
| Unrestricted boosting (interactions) | 0.921 | 0.856 | 0.816 |

Our own published 4-term linear index (AUROC 0.749 at 5 min) does not exceed MAP alone — replicating, in our own model, the deflationary findings reported for commercial predictors [4, 7]. The same nine variables under an additive learner reach 0.919. LepMAP reached 0.821. At 10 and 15 min the additive model *exceeds* unrestricted boosting, indicating that interaction terms overfit.

### 3.3 Additive model: discrimination and calibration

**Table 3.** Additive model, strict scenario, patient-level bootstrap (2 000 replicates). Brier skill is relative to the constant-prevalence predictor.

| Horizon | AUROC (95% CI) | ΔAUROC vs. MAP (95% CI) | Brier | Brier skill |
| --- | --- | --- | --- | --- |
| 5 min | 0.919 (0.909–0.927) | +0.035 (+0.027–+0.043) | 0.0188 | +0.426 |
| 10 min | 0.859 (0.845–0.872) | +0.051 (+0.040–+0.062) | 0.0368 | +0.303 |
| 15 min | 0.825 (0.810–0.839) | +0.055 (+0.043–+0.067) | 0.0527 | +0.247 |

Predicted and observed risk agreed across the full range (lowest decile 0.0 7% predicted vs. 0.12% observed; highest decile 26.2% vs. 25.9%): a 200-fold risk gradient, correctly calibrated.

### 3.4 Operating points

**Table 4.** Operating characteristics of the additive model.

| Horizon | Sens. (Youden) | Spec. (Youden) | PPV | Sens. at 90% spec. |
| --- | --- | --- | --- | --- |
| 5 min | 0.77 | 0.92 | 0.26 | 0.79 |
| 10 min | 0.71 | 0.83 | 0.20 | 0.63 |
| 15 min | 0.69 | 0.79 | 0.21 | 0.55 |

### 3.5 Clinical utility

**Table 5.** False positives avoided per 1 000 patients at equal sensitivity vs. MAP alone (decision-curve analysis). Gains concentrate at low thresholds and long horizons; above *p*_*t*_ = 0.20 the model adds nothing over MAP. We report this honestly as the boundary of clinical utility.

| Threshold $p_t$ | 5 min | 10 min | 15 min |
| --- | --- | --- | --- |
| 0.02 | +104 | +149 | +135 |
| 0.05 | +27 | +84 | +101 |
| 0.10 | +4 | +30 | +56 |
| 0.20 | +0 | +3 | +9 |

### 3.6 Robustness

**Table 6.** Sensitivity analyses, 5-min horizon. he advantage over MAP holds in both monitoring device families and under all grey-zone choices, and is smallest — as selection-bias theory predicts — when no grey zone is excluded (ΔAUROC +0.016).

| Subset / protocol | $n$ | Additive AUROC | MAP AUROC | $\Delta\text{AUROC}$ |
| --- | --- | --- | --- | --- |
| Vigileo/FloTrac | 110 715 | 0.920 | 0.869 | +0.050 |
| EV1000/HemoSphere | 258 253 | 0.919 | 0.888 | +0.031 |
| No grey-zone exclusion | 494 122 | 0.907 | 0.890 | +0.016 |
| Grey zone 65–70 mmHg | 436 357 | 0.900 | 0.869 | +0.032 |
| Grey zone 65–75 mmHg (primary) | 368 968 | 0.919 | 0.884 | +0.035 |

### 3.7 Beyond the grey-zone protocol: a residual leak, and the model’s strongest regime

Auditing our own primary scenario revealed that the field s grey-one protocol retains a residual leak: samples whose current MAP is already below 65 mmHg but has not yet persisted 60 s are neither “already hypotensive’ by the sustained definition nor excluded by the 65− 75 grey one. hey are only 3.2% of strict-scenario samples, yet they carry 62.5% of all positives (event probability 0.67, vs. 0.013 above 75 mmHg): near-tautological predictions that inflate every model’s headline AUROC, ours included. Restricting to unambiguously normotensive samples (MAP *>* 5 mmHg) gives the honest forecasting task — who will cross into sustained hypotension within 5 min from a currently safe pressure. There, the additive model reaches AUROC 0.799 vs. 0.705 for MAP alone: the advantage *triples* to +0.094 . The dynamical trend signals matter most precisely where the current pressure is least informative.

### 3.8 External validation: MOVER (University of California, Irvine)

Following a pre-registered plan (frozen before any external data access: models, score, eligibility, endpoints and gates), we validated the frozen full-cohort additive model, the frozen MAP-only baseline and the frozen points score on the MOVER database [14] (UC Irvine Medical Center; SIS subset, 2015-201 ): a different country, health system, EHR, monitoring workflow and recording cadence (1-min charting vs. 20-s device output; CO and SVV interpolated from staggered nursing chart entries; BIS absent, imputed at its development default). Of 3 candidate patients with both CO and SVV recorded, 39 met the pre-specified eligibility (≥60 concurrent minutes), yielding 9 369 strict-scenario samples (5-min event prevalence 5.6%). A structural property of 1-min cadence is that every sample below 65 mmHg already satisfies the sustained definition, so the strict scenario is automatically the pure-anticipation scenario (current MAP *>* 5 mmHg).

Nothing was refit. he additive model reached AUROC 0. 1 vs. 0.683 for MAP alone: ΔAUROC +0.031 (95% CI +0.003 to +0.063), meeting the pre-registered transfer gate (Δ ≥ 0.02, CI excluding zero), with +0.039 and +0.033 at 10 and 15 min. The magnitude of the advantage is nearly identical to development (+0.035 in VitalDB), despite absolute AUROCs being substantially lower under the coarser cadence, interpolated inputs and sicker case mix - what transfers is the *increment over* MAP, which is the paper s claim. he points score did not meet its transfer gate (AUROC 0.69, Δ +0.01, CI crossing ero): its integer bins are quantised on VitalDB distributions, and we report this as a limitation of the printed sheet rather than recalibrating it post hoc — the continuous model is the transferable artefact.

Two features of the external result deserve emphasis rather than concealment. First, in the unrestricted scenario A — which retains samples already at or below the hypotension threshold — the model does *not* beat MAP (0.853 vs. 0.855). This is the expected consequence of the selection effect this paper documents: when a large share of positives are near-tautological, current MAP is close to unbeatable, and any added value can only appear once those samples are removed. The external cohort therefore reproduces not only our effect but also our methodological argument. Second, the transfer gate was met on 39 patients, so the interval is wide (+0.003 to +0.063) and the point estimate should be read as compatible with a range of effect si es rather than as a precise replication of +0.035.

### 3.9 Shape functions

The nine learned shape functions (Figure 1) are the model. Table 6 reports, for each variable, the exact population-effective range (1st-99th percentile of the per-sample contribution, read directly from the boosted stumps of the frozen model) and the transition zone where the risk gradient concentrates, together with our physiological reading. Figure 1 displays population-averaged (partial-dependence) shapes, which understate contributions confined to sparsely populated regions; the table is the exact account.

**Figure 1.**
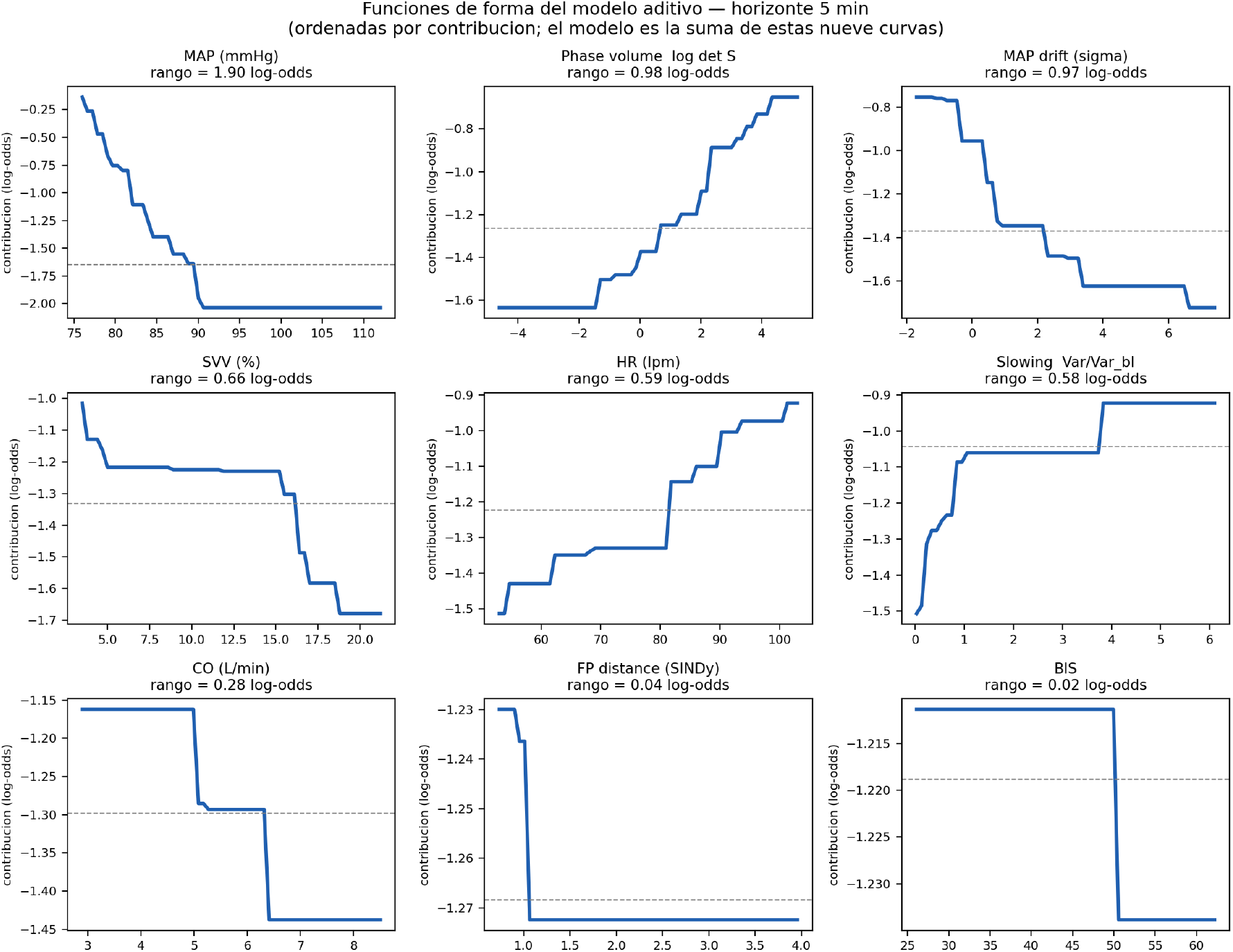
The nine learned shape functions, ordered by log-odds contribution. he model’s prediction is the sum of these curves; there are no interaction terms.

**Figure 2.**
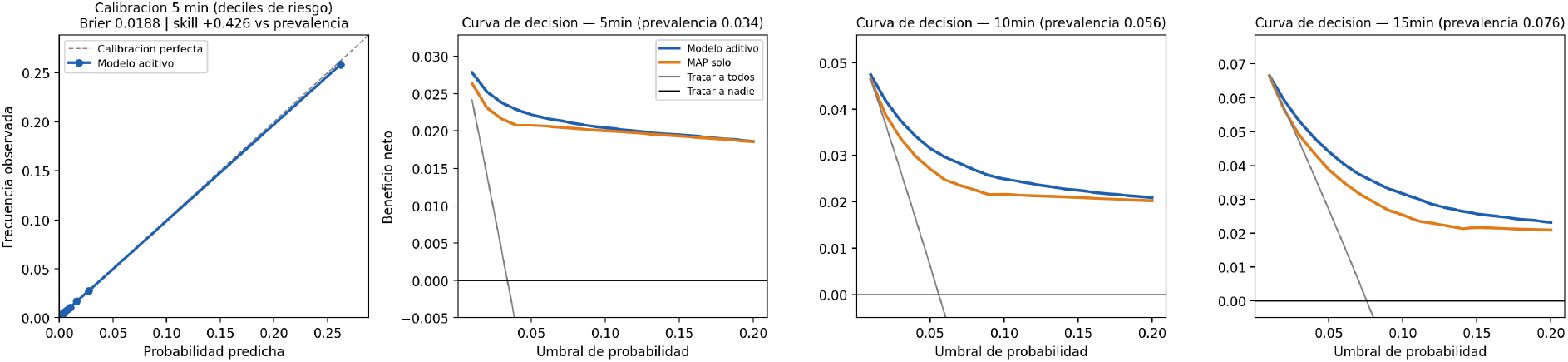
Calibration by risk decile (left) and decision curves for the three horizons (right three panels), strict scenario.

Three findings stand out. First, the MAP shape concentrates its risk gradient across ∼ 79−88 mmHg and saturates above ∼89 mmHg: further pressure elevation confers no additional protection, consistent with a lower autoregulatory margin, and — clinically — with treating pressures in the low-to-mid 80s as already information-rich rather than reassuring. Second, phase-space volume shows flat response below the personal baseline range and a monotone rise once the trajectory expands beyond it (gradient one 0.0-2.3 log det units): threshold behaviour, as bifurcation theory predicts [10]. Third, critical slowing down contributes despite a near-zero *linear* effect size (*d*^*′*^ = 0.04) because its gradient concentrates at variance ratios of 0.15-0.96 - the discriminative change occurs as MAP variance *recovers toward* baseline, saturating once it exceeds it; a linear statistic is blind to a saturating response. We note this reconciliation is post-hoc. The heart-rate gradient is distributed across ∼62-90 bpm with its largest steps above 80 bpm: sympathetic activation as a risk marker overlaps the conventional tachycardia boundary rather than preceding it. BIS shows only a negligible step (at 50) within the population bulk; its large downward step (lighter anaesthesia, lower risk) lies beyond the 95th percentile of observed values, so anaesthetic-depth physiology is visible but population-wise inconsequential. The SINDy-derived fixed-point distance contributes negligibly everywhere (population SD 0.06 log-odds); we report this as an honest negative for equation-discovery features in this setting.

## 4 Discussion

### Answer to the open question

Under the protocol that the field s own critics specified — grey-zone exclusion, no already-hypotensive samples, MAP-based references, patient-level validation [4, 5, 6] — low-frequency haemodynamic trends do add predictive information beyond the current MAP: +0.035 to +0.055 AUROC, with intervals excluding zero, in both device families. The added value is real but bounded: it concentrates at low decision thresholds and longer horizons, and vanishes above *p*_*t*_ = 0.20. A MAP-based alarm remains an excellent default; the additive model refines it where refinement is cheapest — early, low-cost interventions.

### Non-linearity, not variables, was the bottleneck

The ladder localises the information precisely: linear logistic on nine variables retains 28% of the achievable gain at 5 min; allowing each variable a shape function retains 93%; allowing interactions adds nothing further. This has a theoretical reading: early-warning signals of critical transitions are threshold phenomena [10], so imposing linearity on their risk contribution — as fixed-weight indices do, ours included — discards most of their signal.

### The inverted SVV shape is treatment confounding, and we can show it

The learned SVV shape decreases: high SVV predicts *lower* IOH risk, contrary to its mechanistic reading as preload dependence. The data contain the signature of the explanation: in samples with SVV ≥13%, CO rose by +0.18 L/min over the following 15 min and MAP by +0.44 mmHg at 5 min, whereas low-SVV samples showed flat CO (+0.01) and drifting MAP (-0.19). This is the pattern expected if clinicians respond to high SVV with volume: fluid-responsive patients get treated, so high SVV marks *treated* risk. The shape functions are associational displays under standard of care, not causal effects — a caveat that applies to every IOH predictor trained on observational data, including waveform-based ones.

### Relation to waveform deep learning

On this same database, deep networks with high-fidelity waveform access report AUROC up to 0.935 with 14140 patients [8]; the commercial HPI reports 0.95-class discrimination under its original protocol [3]. We do not compete in that regime. Our contribution is orthogonal: 0.05-Hz trends available from any monitor, no waveform licence, evaluated under the bias-resistant protocol that inflates none of these numbers, with full interpretability. Notably, our additive model on trend data approaches the waveform models performance once their evaluation optimism is accounted for — but a same-protocol comparison remains future work.

### An integer points score survives confirmation — if derived exactly

Because the additive model is a sum of per-variable step functions, it admits a points-score representation. Naïve derivations failed: partial-dependence approximation with 4 coarse cuts per variable scored *below* MAP alone (AUROC 0.860 vs. 0.884 ), because the MAP shape concentrates its gradient in a continuous slope (gradient one ∼ 9-88 mmHg, able 6) that coarse bins destroy. Exact extraction changed the outcome: reading the step functions directly from the boosted stumps and pruning them by frequency-weighted dynamic programming to at most 16 rows per variable (113 table rows in total; integer points, 0.05 log-odds per point) lost only ∼0.005 AUROC to quantisation. Under a derivation/confirmation design - 60% of patients for score derivation (6 candidate designs, declared), 40% never touched until a single pre-registered confirmatory evaluation — the score reached AUROC 0.928 vs. 0.89 for MAP alone (Δ +0.03, 95% CI +0.022 to +0.0 6), meeting the frozen gate (Δ ≥ 0.02, CI excluding ero). he deliverable is an APACHE-style lookup sheet evaluable by a monitor or on paper; a -bin pocket card, although it now beats MAP in derivation, was not the confirmed design and is not claimed.

### Why external validation of IOH predictors is scarce, quantified

We attempted to extend validation to MOVER s larger EPIC subset (2017−2022) under a separately pre-registered plan with a declared stopping rule. It could not be done, and the reason is informative. Of 1 85 patients with arterial-line pressure and 912 with pulse-contour cardiac index, 8 1 had both recorded and 207 had temporally overlapping ranges — yet *none* had 60 concurrent minutes of both: within the overlap window the median number of minutes carrying an arterial-pressure entry was zero. In this EHR, invasive pressure is documented occasionally in flowsheets while continuous waveforms live in separate multi-terabyte archives, and advanced haemodynamic monitoring is charted in different care phases from arterial pressure. Per the stopping rule, the analysis was abandoned without computing any performance metric. We report it because it explains a structural fact about this literature: the barrier to externally validating intraoperative haemodynamic predictors is neither the models nor data access, but that the required signals are rarely recorded concurrently and continuously in routine records. This is also why our own external cohort is small.

### Honest negatives

Features derived from sparse equation discovery (SINDy) did not survive: the fixed-point distance ranks eighth of nine (0.04 log-odds), and in a companion analysis a beat-to-beat SINDy anomaly score performed at chance. The early-warning-signal framing survives; the equation-discovery machinery, in this data regime, does not.

### 4.1 Limitations

Single-centre, retrospective data; the label depends on MAP, so MAP-based baselines are structurally favoured (mitigated, not eliminated, by grey-one exclusion); external validation was performed on a small independent cohort (39 patients meeting eligibility in MOVER SIS), so its confidence intervals are wide and a larger external cohort remains desirable; the external data have coarser cadence and interpolated cardiac-output/SVV inputs, so absolute performance there is not comparable to development; net-benefit gains are modest and threshold-dependent; the slowing-down reconciliation is post-hoc; shape functions are associational under standard of care, as the SVV analysis demonstrates; BIS was imputed at 50 where absent. Pooled AUROC mixes between-patient risk stratification with within-patient temporal discrimination: computed within patients (the 6 patients with both classes, weighted by samples), the additive model’s advantage narrows to 0.8 9 vs. 0.869 - real but modest - so part of the pooled advantage reflects identifying higher-risk patients rather than timing events within a patient. For the points score, the derivation/confirmation split preceded all score designs and the confirmatory evaluation was run once; however, the diagnosis that motivated the exact-extraction design drew on earlier full-cohort analyses that included the confirmation patients, so design knowledge is not strictly independent of the confirmation set — a residual, weak dependence that external validation will resolve.

## Data Availability

This study analysed two publicly available databases of routinely collected perioperative data. VitalDB (Seoul National University Hospital) is openly available to all researchers. MOVER (University of California Irvine) is available to researchers who sign its Data Use Agreement, signed by the corresponding author on 19 August 2026. No patient-level data are redistributed by the authors. The complete analysis pipeline, the pre-registered validation plans with every protocol deviation time-stamped before the corresponding results were computed, the frozen models, the out-of-fold predictions, the Lean 4 verification of the points-score quantisation, and the scripts that generate every table in this manuscript are archived at Zenodo, DOI 10.5281/zenodo.22118448.

https://doi.org/10.5281/zenodo.22118448

## Data and code availability

VitalDB is publicly available [9]; MOVER is available to researchers who sign its data use agreement [14]. he pre-registered external-validation plan — models, eligibility, endpoints and gates — was time-stamped before any MOVER data were downloaded and is included in the repository, together with every protocol deviation recorded before results were computed. he complete pipeline — cohort construction, all comparators, pre-registered gates with frozen thresholds, out-of-fold predictions, the frozen model, and the scripts that generate every table in this manuscript — are archived at Zenodo, DOI `10.5281/zenodo.22118448` (concept DOI; always resolves to the latest version). No table in this manuscript was typed by hand; all are generated from the result files whose hashes appear on page 1.

## Use of artificial intelligence

In accordance with ICMJE recommendations, the authors disclose that a large-language-model assistant (Anthropic Claude, accessed through Claude Code) was used extensively during this work: for writing and debugging analysis code, for drafting and revising manuscript text, and as an adversarial reviewer of the authors own analyses. Several errors reported in this manuscript — including label leakage in an earlier labelling scheme, the use of non-causal filtered derivatives, and an effect-size table inherited from a smaller cohort — were identified through that process and are documented in the version history of the public repository. The assistant is not an author. All study design decisions, all pre-registration thresholds, every reported result, and the final text were reviewed and are the sole responsibility of the named authors, who vouch for the integrity and accuracy of the work.

## Supplement S1 - The confirmed points score (design D2a_16)

Derived on the 60% derivation cohort and evaluated once on the held-out 0% (AUROC 0.928 vs. 0.89 for MAP alone). One row per interval; the patient s score is the sum of the nine per-variable point values. Quantisation fidelity to the underlying additive model is formally verified in Lean (*B* = 0.617 log-odds; Methods). Dynamical variables (MAP drift, phase volume, slowing, FP distance) are computed from the 10-min rolling window as defined in Methods.

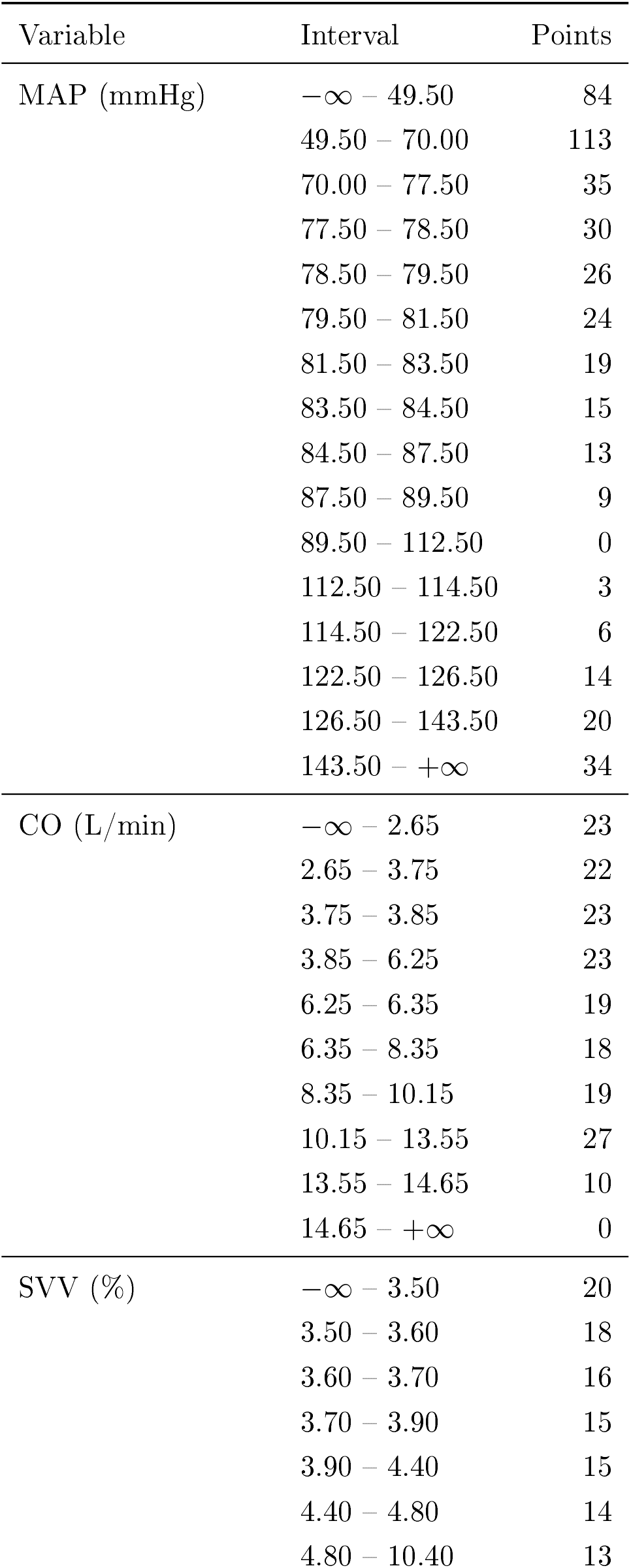
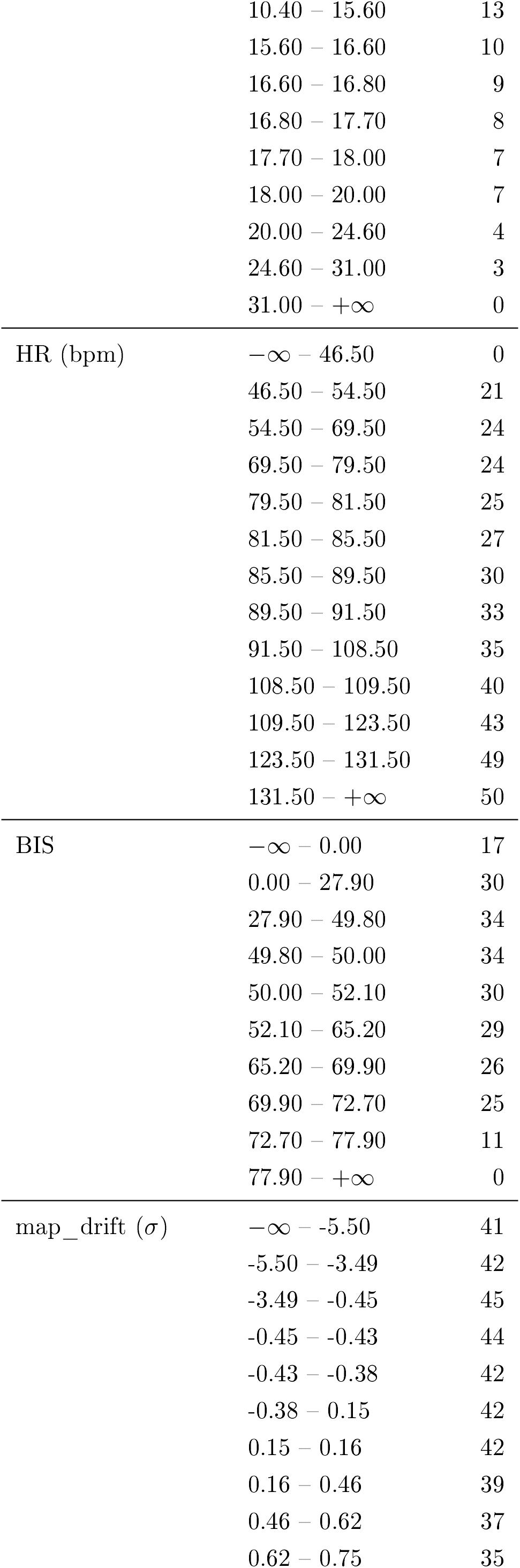
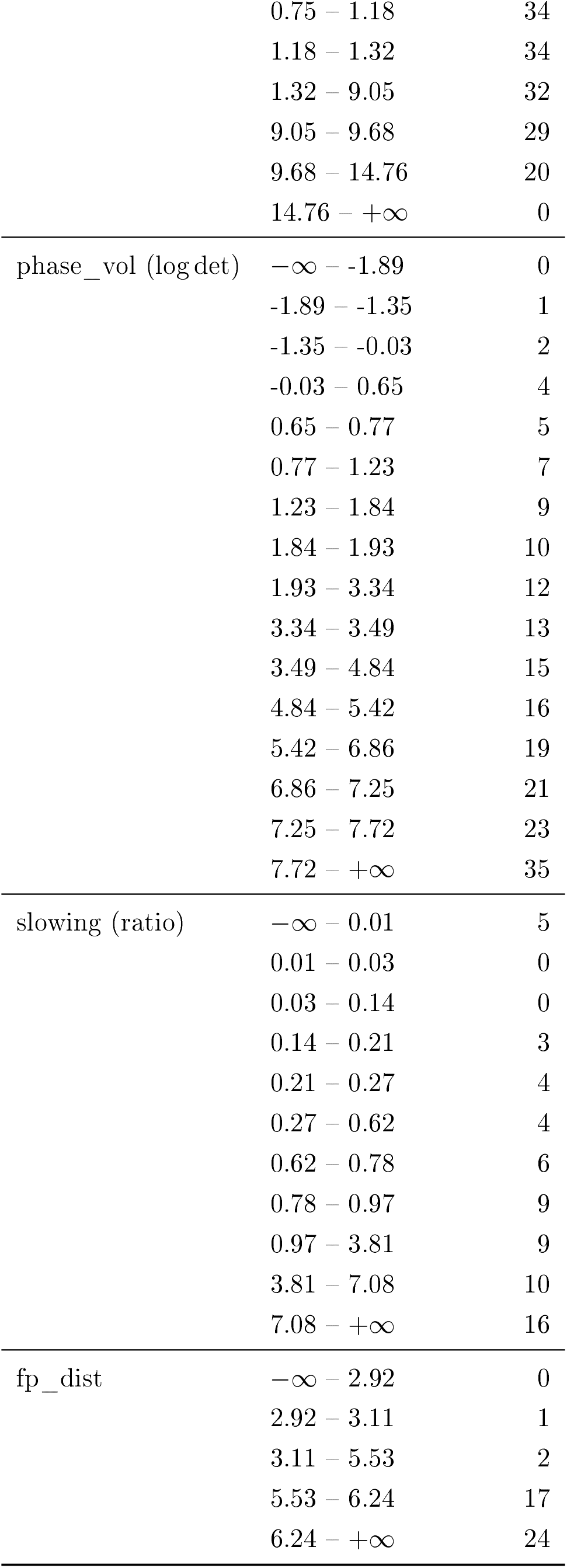

---

**Table 7.**
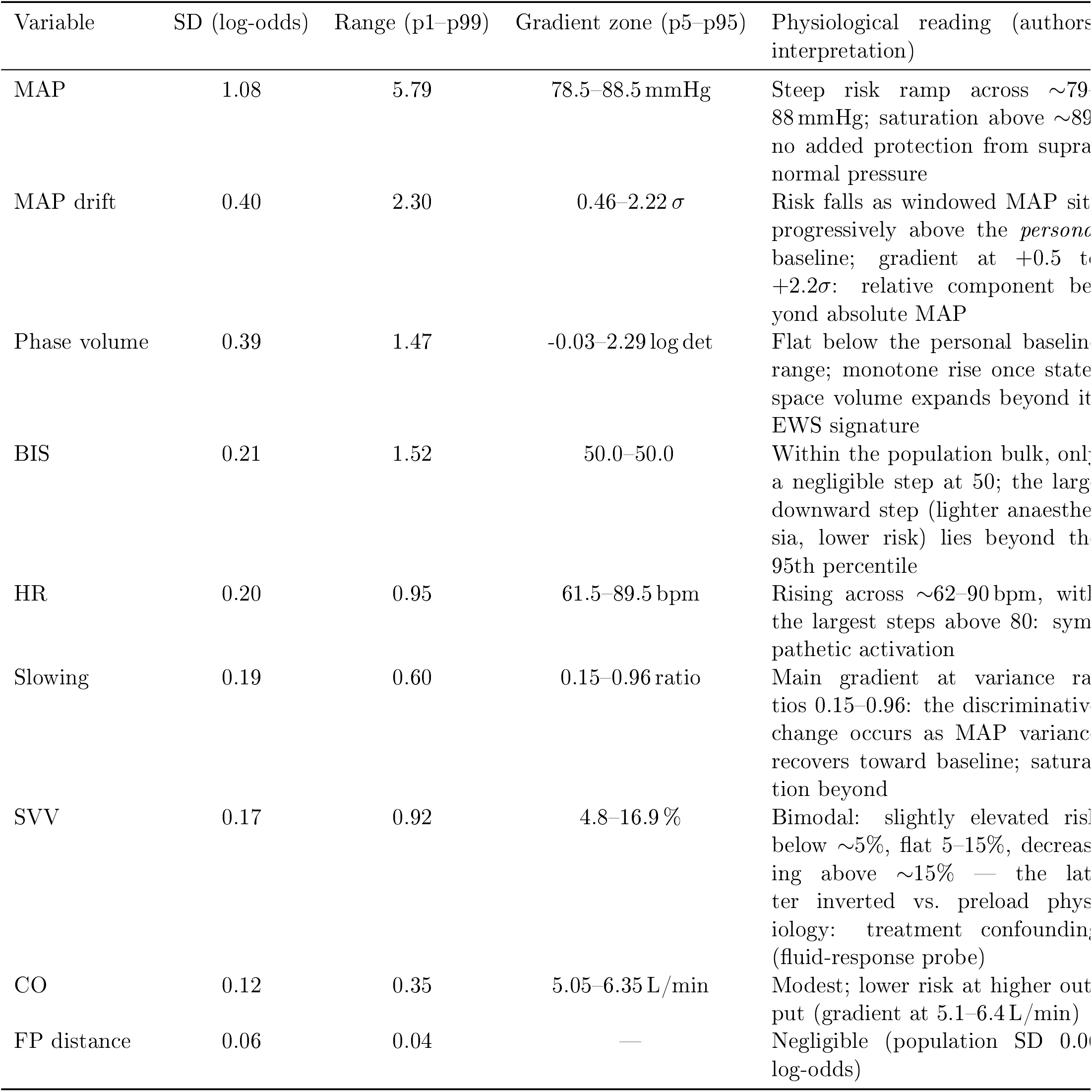
Clinical reading of the shape functions. Numeric columns are computed from the frozen model’s stumps evaluated on the strict-scenario population; the reading column is the authors interpretation (judgement, not computation).

## References

[1] Wesselink EM, Kappen TH, Torn HM, Slooter AJC, van klei WA. Intraoperative hypotension and the risk of postoperative adverse outcomes: a systematic review. Br J Anaesth 2018;121(4): 706–721. doi:10.1016/j.bja.2018.04.036.

[2] Bijker JB, van Klei WA, Kappen TH, van Wolfswinkel L, Moons KGM, Kalkman CJ. Incidence of intraoperative hypotension as a function of the chosen definition. Anesthesiology 2007;107 (2):213–220. doi:10.1097/01.anes.0000270724.40897.8e.

[3] Hatib F, Jian Z, Buddi S, et al. Machine-learning algorithm to predict hypotension based on high-fidelity arterial pressure waveform analysis. Anesthesiology 2018;129(4):663–674. doi:10.1097/ALN.0000000000002300.

[4] Enevoldsen J, Vistisen ST. Performance of the Hypotension Prediction Index may be overestimated due to selection bias. Anesthesiology 2022;137 (3):283–289. doi:10.1097/ALN.0000000000004320.

[5] Vistisen ST, Enevoldsen J. CON: the Hypotension Prediction Index is not a validated predictor of hypotension. Eur J Anaesthesiol 2024;41(2):118–121. doi:10.1097/EJA.0000000000001939.

[6] Jacquet-Lagrèze M, Larue A, Guilherme E, et al. Prediction of intraoperative hypotension from the linear extrapolation of mean arterial pressure. Eur J Anaesthesiol 2022;39(7):574–581. doi:10.1097/EJA.0000000000001693.

[7] Shirmohamadi E, Hosseini Dolama R, Mohammadzadeh N, Ebrahimi N, Ghasemloo N. Intraoperative hypotension prediction in cardiac and noncardiac procedures: is HPI truly worthwhile? A systematic review and meta-analysis. BMC Anesthesiol 2025;25:388. doi:10.1186/s12871-025-03250-4.

[8] Jo YY, Jang JH, Kwon JM, et al. Predicting intraoperative hypotension using deep learning with waveforms of arterial blood pressure, electroencephalogram, and electrocardiogram: retrospective study. PLoS One 2022;17(8):e0272055. doi:10.1371/journal.pone.0272055.

[9] Lee HC, Park Y, Yoon SB, Yang SM, Park D, Jung CW. VitalDB, a high-fidelity multi-parameter vital signs database in surgical patients. Sci Data 2022;9:279. doi:10.1038/s41597-022-01411-5.

[10] Scheffer M, Bascompte J, Brock WA, et al. Early-warning signals for critical transitions. Nature 2009; 461(7260):53–59. doi:10.1038/nature08227.

[11] Vickers AJ, Elkin EB. Decision curve analysis: a novel method for evaluating prediction models. Med Decis Making 2006;26(6):565–574 . doi:10.1177/0272989X06295361.

[12] Olde Rikkert MGM, Dakos V, Buchman TG, et al. Slowing down of recovery as generic risk marker for acute severity transitions in chronic diseases. Crit Care Med 2016;44(3):601–606. doi:10.1097/CCM.0000000000001564.

[13] Collins GS, Moons KGM, Dhiman P, et al. TRIPOD+AI statement: updated guidance for reporting clinical prediction models that use regression or machine learning methods. BMJ 2024;385:e078378. doi:10.1136/bmj-2023-078378.

[14] Samad M, Angel M, Rinehart J, Kanomata Y, Baldi P, Cannesson M. Medical Informatics Operating Room Vitals and Events Repository (MOVER): a public-access operating room database. JAMIA Open 2023;6(4 ):ooad084. doi:10.1093/jamiaopen/ooad084.

